# Transforming Trade for Vaccine Equity: Policy Gaps and Barriers

**DOI:** 10.1101/2024.06.06.24308543

**Authors:** Toby Pepperrell, Meri Koivusalo, Liz Grant, Alison McCallum

## Abstract

The ongoing Pandemic Agreement negotiations illustrate significant gaps in action required to respond effectively to the lessons of the COVID-19 pandemic and make progress towards public health goals, including Sustainable Development Goals (SDGs). The pandemic revealed vaccine equity as a unifying health need, and international trade as a Commercial Determinant of Health. We explored where policy action could reshape trade relationships, identifying recommendations for vaccine equity within stakeholder literature pertaining to Free Trade Agreements (FTAs).

We searched online libraries for stakeholder documents that focused on the interface between FTAs, vaccination, and vaccine equity published between 01/01/2010-31/03/2022. Our analytic framework drew from the rights, regulation, and redistribution (3R) framework, combined with systems analysis, using leverage pointsto categorise recommendations as Technical Mechanisms, Collaborative and Adaptive Mechanisms, or Determinants of Vaccine Equity (DVE). These were then located on a novel systems map to elucidate gaps and actions.

No cohesive strategies for change were identified. Technical proposals were reactive, repetitive, and lacked enforcement mechanisms or incentives. There were significant gaps in the articulation of alternative Collaborative Mechanisms to democratise FTA policymaking processes. The underlying DVE and lack of policy coherence were not addressed. These findings are limited by under-representation of low- and middle-income country authorship in the studies, including in ours, reflecting imbalances in international research and policymaking processes.

Overall, our research shows how the current trade paradigm has produced and sustained vaccine inequity. We propose potential pathways for action but highlight the importance and urgency of more fundamental change in negotiation and implementation of FTAs. New technologies will be crucial for the global response to emerging, neglected, and non-communicable diseases that are vaccine-preventable or -modifiable. Multilateral organisations must, therefore, prioritise the right to health above FTAs , including through TRIPS waivers on Essential Technologies.

## Introduction

Despite globally agreed mechanisms to prioritise global public health over short-term commercial interests and partisan actions by individual governments, vaccine delivery in the COVID-19 pandemic has been inequitable [1]. The Doha agreement and World Trade Organisation (WTO) Trade-Related Aspects of Intellectual Property Rights (TRIPS) flexibilities have proven inadequate in scope and deployment. On May 5, 2023, as the World Health Organisation (WHO) declared the acute pandemic over, low-income countries (LICs) had delivered 5.65-times fewer vaccine doses per adult than high-income countries (HICs) (0.39 versus 2.26; GitHub and World Bank data) [2–4]. It is vital to understand why global access to vaccines has not been achieved.

The role of the Commercial Determinants of Health (CDH) in pandemic preparedness must be examined, including their contribution to vaccine inequity [5,6]. International trade and profit-related movements of goods, people and services played a key role in the emergence and development of the COVID-19 pandemic, including pathways to delivering essential technologies [7]. Vaccines have not been seen as traditional commercially traded products, but as part of international and national public health provision by governments. However, the vaccine industry has changed [8]. Policies and practises arising from Free Trade Agreements (FTAs), including more extensive protection of intellectual property, have affected the manufacture and distribution of vaccines [9], delaying global vaccination. FTAs appear to be having a similar impact on vaccine equity as with new medicines [10].

Vaccines emerge from basic and translational research predominantly funded by the public sector [8]. The expectation that COVID-19 vaccines would be viewed as global public goods (GPGs) was reflected in the resolutions of the 2020 World Health Assembly and UN General Assembly [11, 12]. Instead of acting in global solidarity, however, HIC blocs concentrated vaccine supply, disrupted efforts to pool and distribute vaccines in line with need, and resisted efforts to increase and diversify manufacturing capacity in favour of delayed and inadequate charitable distribution [13]. Vulnerable people and healthcare professionals in low- and middle-income countries (LMICs) remained under-vaccinated, while countries above the charitable income limit found their vaccine supplies delayed, less reliable, and often more expensive than HICs [13].

FTAs promote early market capture of policies related to GPGs at all stages from conception to distribution (Fig 2) with limited attention to the purpose of immunisation as fundamental to the right to health. For example, most FTAs strengthen Intellectual Property (IP) law, protection of trade secrets and commercial interests beyond the WTO minimum (TRIPS-plus agreements) [14]. There is, however, scope for vaccines and vaccination-related services to be considered essential health services and global public goods with long-term benefits including improving planetary health by reducing the risk and consequences of pandemic emergence, and its impact as a driver of pollution and the climate crisis [15].

A planetary health view of vaccine equity considers structural factors that determine vulnerability to exposure to novel pathogens and reinforce inequities in power and resources [16]. Increasing risk of emergence is linked to damaged ecosystems related to biodiversity loss, and animal and human population movements influenced by deregulated trade conditions, especially related to deforestation and agricultural land use [17, 18]

. Meanwhile, severe disease following exposure is associated with poor population health, increased susceptibility to illness, limited infrastructure and intensive strain on the health service with increased use of finite and polluting resources. These are patterned by the Social Determinants of Health (SDH), including, for example, labour conditions, which are shaped by CDH such as FTAs [19, 20]. Populations at greatest risk of significant outbreaks of novel infections are thus dependent on vaccines to limit avoidable harm. Analysis of COVID-19 vaccine equity can, therefore, provide a window on trade as a CDH, and the opportunity to examine planetary health considerations in policy discourse.

We must ask: What can be learned from existing measures and prior global outbreaks? Do trade goals conflict with vaccine equity? What policy incoherencies enable capture by non-health interests? What are the existing narratives for change and who is framing them?

In this study, we sought to assess the associations between vaccine access and FTAs, from basic research to service delivery and the extent to which vaccine equity was considered as a planetary health issue rooted in social and ecological justice.

## Methods

### Literature review

We examined gaps in policy, policy recommendations, and action, with a focus on the role of the WTO and FTAs in the pathways to vaccine equity using the publicly available work of international policymaking bodies and Non-Governmental Organisations (NGOs) with key responsibilities in this area.

We undertook a stakeholder review of the grey literature, complementing an earlier scoping of the peer-reviewed academic literature [21]. We defined stakeholders as organisations with a formal role as policy actors, for example the WTO, SDG custodians, NGOs (international public health bodies, charities, donors, and professional/trade governing bodies with roles in vaccine supply) (Appendix 1).

A starting point for our review was publication by the WHO of the Social Determinants of Health: Conceptual Framework for Action in 2010 [22], which identifies the importance of international trade and industrial policy on the SDH, the role of intersectoral action, and vaccination as one example of interventions that aim to reduce harm from inequitable exposure to vaccine preventable and modifiable disease. We also aimed to incorporate policy learning from Ebola, the H1N1 pandemic and compulsory licensing of anti-HIV medicines.

We searched Policy Commons and online libraries for documents that focused on the interface between FTAs, vaccination, and vaccine equity, enhanced by reference searches and alerts to identify material such as WTO papers becoming publicly available. Following initial screening, we formally searched for English language documents published between 01/01/2010-10/06/2022. We identified additional documents outside this date range from reference searches and publication alerts following the main searches undertaken between 25/05/2022 and 10/06/2022. The documents retrieved formed our dataset (Appendix 2). Appendix 3 includes search terms and PRISMA diagram [23].

Our search window covered initial COVID-19 vaccine distribution during the acute phase of the pandemic, as well as vaccine-related trade policy up to 5 years before the adoption of the SDGs. This decision was taken because SDG 3, particularly Target 3.0.b.01 on universal access to vaccines, provided a formal, global commitment to vaccine equity [24]. It was used as a reference against which we could measure adoption and implementation of policy and practices likely to function as facilitators and barriers to vaccine equity, meeting the UN expectation that trade would be harnessed to meet SDG requirements [25]. We repeated the search on 04/05/2024 to assess whether additional recommendations with transformational potential had emerged to fill gaps identified in our initial review.

We followed the documentary analysis method outlined by Dalglish et al: readying, extracting, analysing, and distilling findings from each document and the relationships between them [26]. Two authors (TP and AKM) skimmed titles and abstracts to determine primary focus, before reviewing in detail to identify policy proposals, actions, and outcomes. We discussed and agreed the findings, fitting them to an analytic framework.

### Development of the analytical framework

Our analytic framework builds on earlier work examining current and potential future approaches to developing sustainable public health and vaccine pathways. We developed the analytical framework by modifying the rights, regulation, and redistribution (3R) model.

### Rights, Regulation, and Redistribution (3R)

The 3R framework focuses on the implications of the legislative framework of international agreements and how these relate to rights, regulation, and redistribution as core elements for social policy and action on the SDH [27]. It is distinct from other global 3R frameworks used in supranational laboratory animal testing and environmental policy for reduce, reuse, recycle.

The 3R model was originally developed as a means of explaining and analysing the impacts of wider global policies on health and social policies [27] and the SDH [28]. It has since been applied to investment agreements to examine how legal transnational frameworks shape policy space for government action [29], and in the trade and health field in the analysis of the implications of trade and investment agreements on health policies.

We applied and adapted the 3R framework for the purpose of this review (Fig 1) [28]. It is attractive methodologically because it enables focus on the international agreements and bridges the gap between examination of the social determinants of health (including equitable health systems) and the CDH (of which trade and investment agreements are one) [30], particularly the role of transnational actors as ‘vectors of disease’ [31]. The 3R framework also focuses on ways in which governments and bodies charged with multilateral governance can act to secure the right to health, including access to medicines and healthcare, for example in relation the role of investment agreements and investment protection [29] .

**Fig 1:**
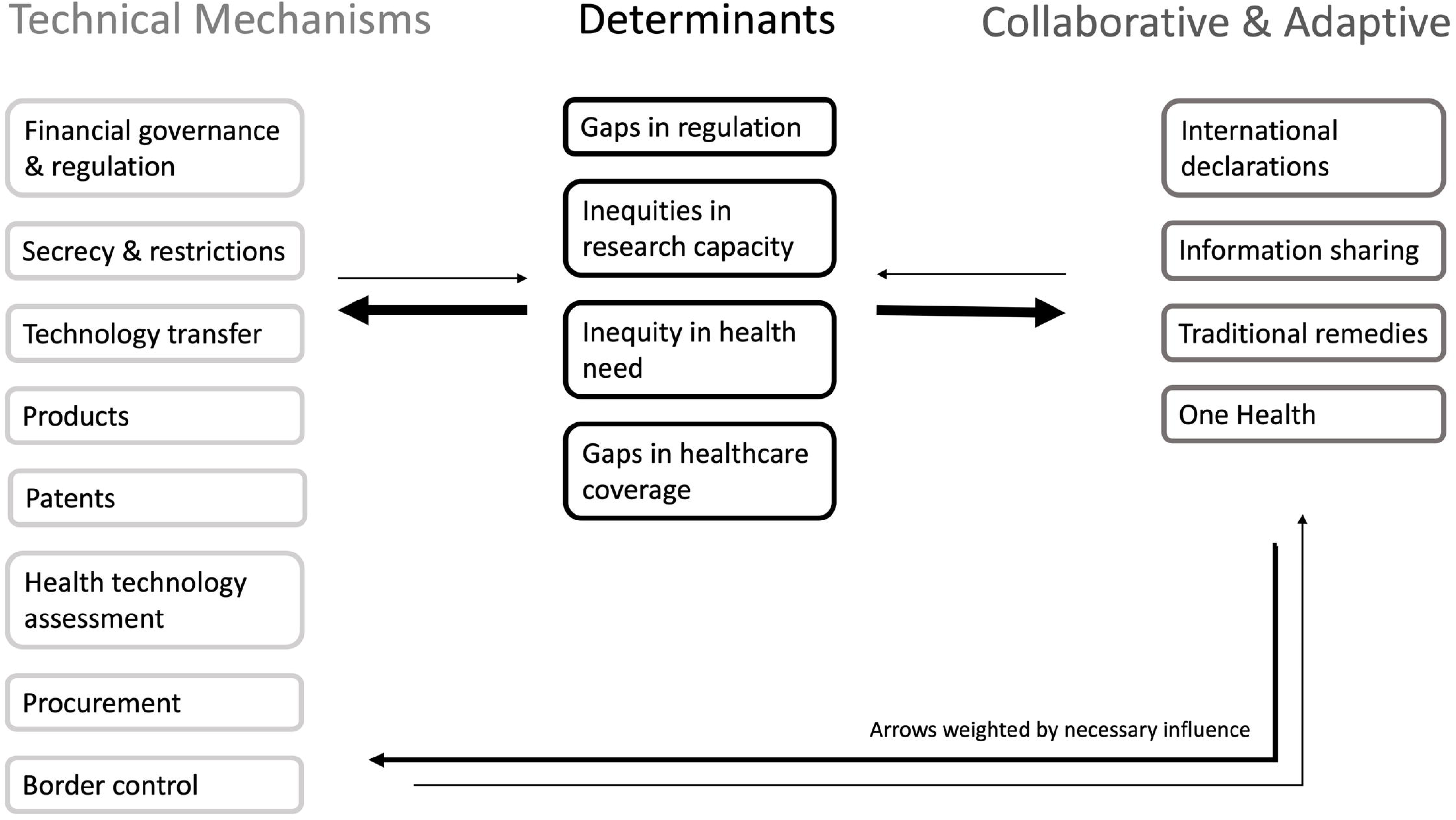
Analytical framework adapted from the Rights, Regulation and Redistribution Framework

**Fig 2:**
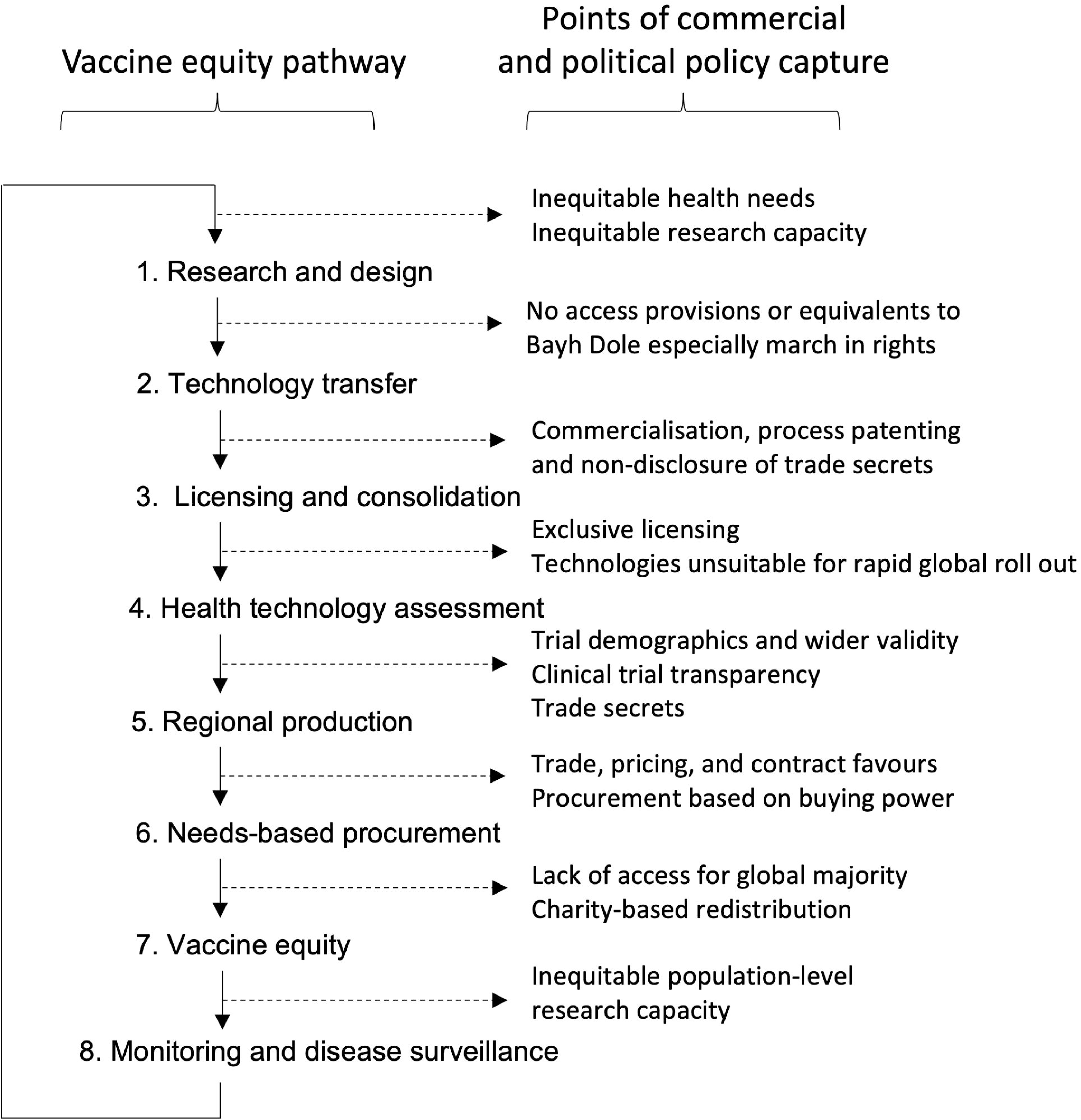
Systems map showing points of policy capture from vaccine research and design through to marketisation and distribution.

### Meadows’ systems analysis

We combined the 3R framework with Meadows’ systems analysis, which has previously been applied to public health issues [32], and the implications of commercial policy upon public health policy [33]. Our current usage reflects this wider application. Trade agreements can be understood as systems as well as means of shaping systems – allocation of rights and regulations with implications for redistribution and equity [34, 35]. Here, leverage points in the system relate to potential levers of change that were initially articulated regarding concerns about trade policy and WTO as a system [36, 37].

### Application of the analytical framework

We looked specifically at factors that would affect enforcement of the right to health and impact multilateral regulation for equity in vaccine development and distribution, and their points of impact on the system. These factors were then mapped onto the analytic framework. The overarching categories can be considered at three positions along Meadows’ leverage points to intervene in a system [36], grouped as: Determinants of Vaccine Equity (DVE), Technical Mechanisms, and Collaboration and Adaptation around the global free trade environment. Subcategories from the 3R framework were expanded as themes emerged in analysis.

### Definition of transformative potential

The criteria for judgment on transformative potential was made on the grounds of the differential potential of leverage points, in line with consideration of proximate and root causes of health concerns [38]. Technical Mechanisms are proximal and visible, addressing specific gaps without effecting deep or sustainable transformative change; Collaborative approaches, shared goals, professional and organisational responsibilities, can enable greater cohesion, but do not necessarily address determinants. Alone, they are rarely transformative but can be its starting point. Determinants are underlying causes from which pervasive political and commercial health effects emerge, Attention to determinants is thus most likely to be transformative [39]. Potentially transformational policy interventions were those designed to shift the dial on the fundamental causes of vaccine inequity and enforcement of right to health obligations. This included action to reduce imbalances in power, resources and money on a transnational scale. Other potentially transformative interventions encompassed action to address one or more of the persistent gaps in governance that sit at the interface between trade and investment agreements, public health and health equity. Examples include capture of decision-making by high-income countries and corporations, weak accountability and scrutiny of the impact of decisions on equity, institutional stickiness that sustains current inequities, and ways of doing and being that constrain rather than expand policy space for health [40].

We discussed the findings first as broad themes under each category and then examined the subcategories, focusing on advancing vaccine equity at specific points in the causal process (Fig 2). This allowed us to interrogate which recommendations could be transformative and identify gaps.

## Results

We screened 10,000 documents by abstract and title, 115 papers and reports met our eligibility criteria and underwent full text review (Appendix 2). Forty papers were published in the pre-SDG period from 2010-15, 24 after the adoption of the SDGs, and 51 in the acute phase of the pandemic. Sixty-nine were excluded as they contained no action points (n=25), provided only basic information (n=18), provided no health (n=9), or trade policy (n=8) commentary, full text was inaccessible (n=6), or they were not international (n=3) (Appendix 3). Of the 46 documents included, only 12 came from stakeholders in the Global South (Appendix 1). Stakeholder references from the repeated search were not included in the dataset, as they did not pertain to the acute phase of the pandemic or reveal any novel recommendations, but search results are available on request.

We identified 267 recommendations likely to influence vaccine equity. Those that could enable significant vaccine progress towards SDG 3 were considered potentially transformational (Table 1). Technical Mechanisms constituted 152/267 (56.9%) proposals, of which 12/152 (7.9%) were considered potentially transformative, 48/267 (18.0%) focused on Collaborative and Adaptive Mechanisms, of which 8/48 (16.7%) were transformative, while 67/267 (25.1%) addressed DVE, with 9/67 (13.4%) transformative (Table 1). Our updated search identified no new transformational recommendations, though additional examples of Technical and Collaborative mechanisms were identified for points a.ii, c.ii, d.ii, g.i, i, o.iii in Table 1 [41–48].

**Table 1:** recommendations in the available policy literature around (a.) Technical and (b.) Collaborative & Adaptive mechanisms to improve vaccine equity, and (c.) the Determinants of Vaccine Equity (DVE). *Links to Dataset Appendix 2, not bibliography.

| Category |  | Recommendations |  |
| --- | --- | --- | --- |
| a. Subcategory |  | Number (/267) | Potentially transformational (/29) |
| i) Themes in findings |  |  |  |
| Technical mechanisms |  | 152 | 12 |
| a. | Patents<br>i) FTA consultation with WIPO, WTO and WHO on public health measures<br>ii) TRIPS modifications and TRIPS-plus flexibilities<br>iii) Voluntary and compulsory licensing mechanisms<br>iv) Emergency measures | 39 | a.ii.iii.iv.1 Patent waiver enacted during pandemic for vaccine technologies and components, vaccines, and vaccine-related products, including know-how and two-way education programmes (2*, 7*, 28*, 43*, 61*, 88*) |
| b. | Financial regulation and governance<br>i) Regulation of FTAs<br>ii) Health technology markets and foreign investment<br>iii) TRIPS flexibilities and compulsory licensing, article 31<br>iv) Direct trade interventions<br>v) Health system strengthening methodology | 36 | b.i.1 Primacy of public health goals and actions in FTA negotiations (2*, 8*, 38*, 42*)<br>b.i.2 Open investigation of effects of trade openness on deforestation and zoonotic risk to be moderated by land rights and financial guidelines (89*)<br>b.ii.1 Transparency on international investment rules as part of a multilateral governance framework alongside SDGs (72*)<br>b.iii.1 Open public discourse on technocratic and political barriers to TRIPS flexibilities (38*, 42*, 46*, 55*, 58*)<br>b.v.1 Debt crisis solutions via United Nations brokering in recognition of consequences of debt on public health (66*) |
| c. | Products<br>i) International harmonisation and clarity<br>ii) Vaccine inputs and global supply chain<br>iii) Wider production capacity<br>iv) Charitable interventions<br>v) Emergency measures | 19 | c.i.ii.iii.iv.v.1 Experimental policies to address barriers to supply diversification, including pre-definition of emergency supply chain procurement procedures, requiring preparatory mitigation of supply chain risks, establishing new reporting and monitoring methods, moving on from just-in-time procurement practices, and reducing geographic distance and fragmentation by incentivising regional or nearshore procurement, using a multiple hub approach (20*, 27*, 93*) |
| d. | Procurement<br>i) Self-determination: procurement policy that reflects national priorities<br>ii) Multilaterally mediated pooled procurement process for all LMICs<br>iii) Competition and equity<br>iv) Transparency | 16 | d.ii.1 More extensive application and use of pooled procurement mechanisms (7*, 8*, 20*, 43*, 69*, 88*, 105*)<br>d.i.ii.iii.iv.1 New approaches to procurement by prequalification based on potential harms of lack of rapid and equitable vaccine access (8*) |
| e. | Health technology assessment<br>i) International harmonisation and clarity<br>ii) Transferability<br>iii) Clinical trial data<br>iv) Cost-benefit approach to prequalification | 15 | e.i.iii.iii.iv.1 International collaborative approach to health technology assessment, and agreed criteria for rapid assessment and approval in any nation (5*, 6*, 8*, 35*, 44*, 74*) |
| f. | Border control<br>i) Import-export restrictions and tariffs<br>ii) Bottlenecks<br>iii) Paperless trade<br>iv) Long-term agreement and definitions | 14 | .. |
| g. | Technology transfer<br>i) Access included in governance of privatisation of public research (Bayh-Dole equivalent public tasks for private corporations)<br>ii) Pooled access initiatives require engagement at R&D phase | 7 | g.i.1 Public health criteria strengthening: Bayh Dole equivalents (legislation to ease commercialisation of high-priority products resulting from public research) to have ‘march in’ rights if companies not enabling products to be made or distributed at appropriate scale to meet public health needs (104*)<br>g.i.2 Policies require equitable access provision at the point of public-to-private technology transfer (2*) |
| h. | Secrecy and restrictions<br>i) Intellectual Property law | 6 | .. |
|  | **Collaborative and adaptive mechanisms** | 48 | 8 |
| i. | Information sharing and transparency<br>i) Data and know-how within and between healthcare systems<br>ii) Cost transparency for negotiation capacity | 21 | i.i.ii.1 Interoperable data sharing systems (8*)<br>i.i.ii.2 Designing transparency into all practices from research through development, including funding and conflict of interest (6*, 35*) |
| j. | International declarations<br>i) Balance of corporate compared to community rights and obligations<br>ii) Revising outdated or dysfunctional agreements<br>iii) Novel agreements | 17 | j.i.ii.1 Broad vaccine delivery partnership boosting microplanning through advocacy and political engagement within UN – integrated ground level teams associated with regional and global partners (111*)<br>j.iii.1 Reshape minimum requirements of Medical Innovation Prize Fund and similar mechanisms– incentivise strategic global health benefit at generic price (104*) |
| k. | One Health<br>i) Universal Healthcare<br>ii) Vaccine programmes<br>iii) Environmental | 8 | k.iii.1 Common minimum environmental standards to be agreed for FTAs, with civil society involvement (111*) |
| l. | Traditional knowledge<br>i) Registry and recording<br>ii) Patentability and protection | 2 | l.i.ii.1 Enforceable rights for custodians of traditional knowledge to protect knowledge streams and ensure benefit sharing from resultant innovations (48*)<br>l.i.1 Essential R&D into fostering R&D potential and knowledge-based infrastructure led by discriminated populations in LMICs (104*) |
|  | **Determinants of Vaccine Equity (DVE)** | **67** | **9** |
| m. | Gaps in regulation<br>i) Empirical policy debate and legislation<br>ii) Borders<br>iii) Pricing<br>iv) Safety, pharmacovigilance, and ethics<br>v) Corporate and professional conduct related to vaccination | 28 | m.i.1 Address imbalance in corporate vs planetary interests by moving from best endeavour e.g. labour, environment, agriculture, public health requirements into hard law commitments similar to e.g. finance, capital investment, IP rights in enforceability (111*)<br>m.i.iv.v.1 Design pharmaceutical education curricula and care plans to meet local needs from practice level assessment and not just minimum international guidelines (43*) |
| n. | Inequities in research capacity<br>i) Regulatory<br>ii) Innovation<br>iii) Validity<br>iv) Access to medicines<br>v) Transparency | 21 | n.i.ii.iii.iv.v.1 Law to support local R&D and enshrine regulation of major corporations undertaking R&D and production in diverse settings (104*)<br>n.i.ii.iii.iv.v.2 National self-definition of R&D priorities before externally imposed intergovernmental definition (43*)<br>n.iii.iv.1 Large-platform trials to allow for experimental therapies to be added and dropped adaptively in order to create a wider and more rapidly evolving evidence base for novel and repurposed therapeutics (116*) |
| o. | Inequity in health need and access<br>i) Rights-based financial support<br>ii) Fiscal justice<br>iii) Addressing harms and gaps in right to health | 11 | o.i.ii.1 Nuanced financial framework responding specifically and appropriately to socially determined health needs in a rights-based manner, rather than average national income (46*, 53*)<br>o.ii.iii.1 Open discourse and action on adverse impacts of debt repayments, especially interest above initial loan, on health systems and pandemic response (66*)<br>o.i.iii.1 Structural provision for women's rights organisations to mitigate the gendered impacts of the pandemic and vaccine inequity (66*) |
| p. | Gaps in healthcare coverage<br>i) Funding wastage<br>ii) Healthcare worker movement and rights | 7 | p.i.1 Sustainable funding of prevention, treatment, and care pathways through agreements around global public goods or generic provision, avoiding excessive spending and reliance on specific proprietary technologies that crowd out other aspects of service provision (104*)<br>p.ii.1 Special mutual recognition of access to healthcare for migrant healthcare workers, and in free movement agreements (88*, 103*) |

### Thematic Analysis

We drew out the processes involved in vaccine development, production, distribution, and service delivery, and identified where FTAs and trade-related policies and procedures had the potential to facilitate or constrain efforts to progress vaccine equity. Overall, potentially transformational recommendations either target points of commercial and political policy capture (Fig 2) or aim to transform the governance sphere to influence the way in which decisions are made.

### Technical Mechanisms

Development and application of technical mechanisms that limit or facilitate access to vaccines dominated the policy discourse. Technical recommendations focused on addressing vaccine inequity post-policy capture (Fig 2). Patents, supply chain and borders issues dominated (Table 1, a.-d., f.), tending to provide workarounds to mitigate short term harm rather than transformation. As our dataset spanned a 20 year period, it is clear that this short-termism is not an isolated phenomenon of the pandemic period where policies had to be pushed through quickly.

Almost two-thirds of regional FTAs include TRIPS-plus agreements [49]; one vaccine can entail multiple patents and trade secrets covering essential technologies and processes [50]. Without access provisions at a public-private technology transfer stage, new FTAs and TRIPS-plus agreements afford market exclusivity to the few companies that own patents, proprietary technology, and trade secrets for periods that extend beyond the acute phase of an outbreak or pandemic. Few stakeholders acknowledged the importance of early intervention to support public development, prevent or limit exclusive licensing (Fig 2, 1.-3.), and assure adequate governance to prevent market domination and excessive profit-taking (Table 1, g., h., i., n.iv.v). Without effective interventions, supply is capped. In addition, few countries produce vaccines, so most governments have limited scope to use domestic legislation to address emerging inequities, ensure affordability, or investment in infrastructure development.

Documentary analysis repeatedly identified Article 31 on TRIPS flexibilities [51]. Compulsory licensing is designed to combat TRIPS-related inequity of access to medicines, but complexity, potential costs, and lengthy timescales have limited its use (b.iii.1) [52,53]. Concern about the risk of trade and non-trade sanctions has limited repurposing of existing facilities and reverse engineering of vaccines (Table 1, b.iii.1, c.i.ii.iii.iv.v.1) [54]. Significant effort has been expended on complex negotiations and workarounds, while the WTO has recognised that TRIPS flexibilities were designed to address national rather than global emergencies [55]. To effect responsive vaccination to curtail a polio outbreak in Israel, the manufacturer waived the patent voluntarily, enabling local production [56]. The original compulsory licensing framework relied on exceptional conditions and, when designed, did not anticipate the range of behaviours of companies or vaccine-producing trading blocs that now distort the relationship between supply and need [53]. Few stakeholders addressed the relatively weak measures available to address failures to protect public health. Legal measures to formalise research ethics and public protections in law were key themes despite receiving little public attention.

### Collaborative and Adaptive Mechanisms

We identified calls for open communication and information sharing with interested parties (Table 1, i.). Among the best-established examples are those for globally sharing intelligence, tissue, data, and expertise to support horizon-scanning and syndromic surveillance for emerging threats to health for vaccine preventable and modifiable diseases [57]. These efforts sit alongside advocacy for clinical trial transparency, action on price negotiations, epidemiological mapping and supporting infrastructure [58,59]. However, Collaborative Mechanisms should also provide alternative means of resolving trade related issues related to vaccine equity. Significant gaps and inconsistencies impede this possibility [60]. In addition, while some grassroots and NGO efforts addressed supply chain issues, the role envisaged for other than market-based actors or activities, including governments, was minimal.

Collaborative and adaptive approaches should provide enabling mechanisms for public health FTA exemptions as a minimum, as attempted by the Medicines Patent Pool (MPP), a WHO platform for pharmaceutical companies to negotiate voluntary licences [61]. Accessing technologies and know-how through the MPP, generic firms can begin drug development, which is associated with lower costs, greater diversity of clinical trial participation, and greater HTA approval rates [62]. However, such efforts remain context and topic specific.

Without a systems approach, positive examples remain largely invisible to wider FTA decision-making. Equity must be upheld as a collaborative process and outcome, but we found public health measures reduced to specific interventions, reflecting hard-won, case-by-case global health diplomacy rather than progress towards system redesign. We found no proposals for community or grassroots representation in decision-making processes from the bodies responsible for multilateral governance.

### Determinants of Vaccine Equity (DVE)

There was no clear pathway to deliver vaccine equity in line with the requirement for universal access to vaccines. The Doha Declaration on the TRIPS agreement and Public Health and subsequent amendments allow for measures to address public health problems, including through vaccination [53,63]. However, we found limited evidence of attention to the structural, systemic, and institutional barriers to vaccine equity associated with FTAs including the trade-related issues that complicated the response to Ebola [64,65]. After limited progress towards a more comprehensive pandemic waiver in WTO [66], equity and access questions are ongoing within the Pandemic Agreement negotiations and were raised in the revision of International Health Regulations (IHR) [67]. While international negotiations have the potential to change the balance of global governance, the completion of the IHR negotiations and the Pandemic Agreement discussions in the WHO do not indicate that these would bring a major change in relation to the current WTO framework of governance [68]. Pandemic Agreement negotiations have been through many rounds and are expected to continue until May 2025. In theory, a Pandemic Agreement could have provided leverage for governments to introduce safeguards and regulation to ensure rights of governments to secure access to diagnostics and vaccines for the purpose of pandemic control and equity in access to vaccines, including with respect to pathogen access and benefit sharing systems, financing, and technology transfer. However, while some progress has been achieved [69], this may be more limited than hoped for with respect to the Pandemic Agreement by G77 and nongovernmental actors [70,71].

### Analysis of Specific Recommendations

Technical recommendations frequently referenced procompetitive corporate governance (Table 1, a.-b.). Processes such as patent thickets and evergreening complicate an already resource intensive pathway for LMICs to access vaccines within WTO rules [41]. Corporate rights also dominated policy discourse. Narrowly drawn recommendations may illustrate a deliberately incremental approach favoured by some authors but there was little evidence of a strategic plan for vaccine equity in the stakeholder literature [72]. Just 6/152 recommendations (3.9%) addressed secrecy and restrictions (h.), and 7/152 (4.6%) technology transfer (g.), both crucial to vaccine equity, blocking or undoing policy capture at phases 1, 2, and 3 of the vaccine equity pathway (Fig 2).

Twelve Technical proposals were potentially transformational (Table 1). One, patent waiver (a.ii.iii.iv.1), directly addressed patents, trade secrets and non-patent related IP (a., h.). Propositions included a multilateral investment framework compatible with the SDGs (b.ii.2), breaking down barriers to employing TRIPS flexibilities (b.iii.1), and equitable vaccine research and production processes with ‘march in’ rights where products are not being made or distributed at scale to meet public health needs (g.i.1-2).

Collaborative recommendations focused largely on pre-existing declarations (17/48, 35.4%) (j.) such as implementation of TRIPS amendments or mechanisms designed to increase transparency (21/48, 45.8%) (i.) [51]. These often fall back on best endeavours rather than enforceable requirements or agreements formalising collective commitments, intelligence, and action. Potential catalysts for transformation included more comprehensive commitments to transparency and knowledge sharing (i.i.ii.1-2), alternative vaccine delivery partnerships (j.i.ii.1) and unlocking LMICs’ R&D potential. (l.i.1), which all pertain to the redistribution of power necessary for greater transformation.

Recommendations considering the social, political, and commercial DVE as a subset of health equity – other than increasing average national income – were infrequent. Discourse on the determinants of health revolved around gaps in (financial) regulation and increasing the potential of LMICs to undertake innovative commercial health technology research (49/67, 73.1%) (m.-n.) rather than attention to rights, redistribution, or sustainability (o-p.). Gaps in healthcare provision, access to care (11/67, 16.4%) (o.) and underlying causes of health and healthcare inequities that manifest as barriers to vaccine equity were overlooked (7/67, 10.4%) (p.). In the Determinants category, potentially transformational recommendations included calls to strengthen legislation around planetary health versus corporate interests (m.i.1), tailored financial support to address the SDH at community (o.i.ii.1) and macro levels, including addressing the impacts of debt repayments (o.ii.iii.1). These factors have the potential for impact at multiple points on the pathway to vaccine equity including research and development, technology-based transfer and needs-based procurement. They thus demonstrate the opportunities for transformation by taking a systems approach and using this to apply multiple levers, each of which addresses barriers to vaccine equity.

### Inter-related Nature of Recommendations

Not enough attention has been paid to the relationships between individual recommendations. Technical Mechanisms often depended on DVE, for example releasing resources for health system strengthening by revoking or minimising the impact of debt repayments (b.v.1, o.ii.iii.1), but there is currently no clear Collaborative bridge for mobilisation. An example of this is seen with the recommendation for national self-determination of research and healthcare goals in LMICs (e.ii.iii.iv.1, l.i.1, n.i.ii.iii.iv.v.1-2).

## Discussion

There was insufficient recognition of FTA impact on vaccine equity in the international stakeholder literature. Incremental fixes such as TRIPS flexibilities and case by case approaches such as compulsory licensing mechanisms [73] were not linked to new forms of collaboration or solution-building (Table 1, i.-l.). Siloed technical solutions overwhelmed efforts to address the building blocks of vaccine equity such as reforming undemocratic decision-making (Table 1, a.ii.iii.iv.1, b.iii.1), enabling technology transfer (g.i.1) and addressing barriers including patents and trade secrets (a.ii.iii.iv.1, i.i.ii.2). While Article 7 in the TRIPS agreement calls for balance between property rights holders and users, this is contested terrain as the high costs of new medicines and delivery systems result in health system pressures that limit access and crowd out investment elsewhere in health and care. Though the TRIPS agreement was not initially the main barrier to COVID-19 vaccine access, it has increasingly constrained movement to address inequity in access and affordability [74]. This current review aimed to highlight current gaps and opportunities for change.

Vaccine equity could have been designed into global pandemic preparedness and response based on learning from the Ebola vaccine and antiretrovirals for HIV [65, 53], but efforts were diverted by a best endeavour framing of public health needs that lacks the enforceability that currently exists uphold the rights of corporations. The European Union (EU), the UK, and the US were able to veto the COVID-19 technologies TRIPS waiver despite support from around 100 nations and calls for international cohesion from WHO, WTO and WIPO leadership [75]. As new WTO regulations typically require consensus, countries with stronger negotiating positions can block transformational proposals [76], limiting progress towards vaccine equity. Trade-offs and compromises across different areas of negotiation can also undermine improvements. WTO and WIPO are thus unlikely to be able to support transformative measures to enhance vaccine equity but will be bound to expanded and strengthened global agreements [49].

### Addressing Gaps in the Current Approach to Addressing Vaccine Equity

Technical Mechanisms are vital tools that can enable the introduction of specific interventions that address barriers or enable vaccine equity. However, they formed no coherent strategy in the policy discourse, a minimum requirement for pandemic preparedness. Discussions on co-created models of financial support (o.i.ii.1) were overshadowed by those imposed by HICs and multilateral organisations, particularly GDP (Gross Domestic Product) and World Bank national income category as proxies for resource availability. MSF (Médecins Sans Frontières) Access reports illustrate that LMICs are subject to cliff edges in funding from international development organisations like Gavi when national income or GDP reaches an externally imposed threshold [77]. There has been little recognition that modelling and pricing processes do not take need, purchasing power parity or affordability into account. Rather than assuring the right to health, the global COVID-19 vaccine programme has been directed by growth-oriented FTA economics that simplifies complex geopolitics. There were no proposals for more inclusive shaping of international trade beyond the existing WTO regulation of FTAs. Instead, energy had to be directed towards resolving preventable issues like vaccine dumping.

Costa Rica’s proposal for a global technology and IP pool in March 2020 and Eswatini, India, Kenya, and South Africa’s proposal for a TRIPS waiver were important interventions that were rebutted [78,11]. Instead, underdeveloped Collaborative Mechanisms and limited multilateral governance undermined the ACT-A and COVAX collaborations and the additional emergency measures proposed. This failure is reflected in the IHR and Pandemic Agreement negotiations as LMICs advocacy for global equity has received significant pushback [79,80]. An enforceable global IP pool or TRIPS+ waiver including action regarding, for example, trade secrets or measures to limit profiteering, would have facilitated greater vaccine equity and informed wider corporate regulation.

We propose that access goals should be enshrined in law, supporting progress towards SDG 3 commitments, including universal access to vaccines. Existing mechanisms requiring corporations to fulfil public tasks before allowing the exclusive licensing of essential medicines and technologies that limits their distributive potential in health emergencies, could be built on. This would extend the disaster prevention and major incident response requirements placed on certain industries to pandemics [81]. IP regulations must ensure that public health measures can be enacted rapidly, dismantling patents or trade secrets as barriers. To build on the success of the pre-prepared protocols and mechanisms for rapid resourcing and implementation of SARS-CoV-2 vaccine trials, there must be pre-defined conditions and methods for waiving patents and trade secrets on pandemic products, failing removal from TRIPS coverage [82]. While these issues, including benefit-sharing, are included in the Pandemic Agreement, its scope indicates limited progress [79].

Vaccine equity requires a focus on collaboration over competition. Corporate commitments to transparency may be welcome first steps but will not deliver the improvements in the DVE or lower vaccine need; they have previously been used to argue that deeper change to IP and trade secrets is unnecessary [83]. This implies that the transformative potential of cooperative action and non-for-profit collaboration has not been considered. Without greater connection between populations, developments like the MPP cannot function as desired. The lack of an overarching strategic approach means that exclusion and inequity are baked into current FTA governance. For equity to be integral to pandemic preparedness, decision-making must centre independent regional, NGO, and grassroots civil society, currently excluded from closed-door negotiations.

Our stakeholder review found that power imbalances, postcolonial trade justice and human rights obligations, were under-recognised [84]. LMIC voices, particularly in-country NGOs, and advocacy bodies, were barely present; we identified only 12 policy documents from the Global South. Without a critical lens on how policymaking processes contribute to the determinants of health, opportunities for vaccine equity were missed throughout the pandemic. For example, available mRNA vaccines had exacting cold chain requirements.

Community-based LMIC-led innovation could reduce barriers to local production, energy- and resource-dependent delivery, and hesitancy [64,65].

Action to address the flaws and limitations of current multilateral governance mechanisms is required, particularly in relation to the roles of the WTO and the WHO. Table 2 gives our synthesis of priorities for action. Trade is a tool, not an outcome, and public health must be consistently central to FTA negotiations, with enforceable definitions of compliance with the right to health as a corporate obligation rather than a task-specific, incentivised, discretionary mechanism.

**Table 2:** Priorities for action, building on recommendations from the stakeholder review (Appendix 2) and addressing gaps in the pathway to vaccine equity (Fig 2)

| Aims | Objectives to address policy incoherence | Immediate steps |
| --- | --- | --- |
| **Overarching**<br>Develop a strategic plan for vaccine equity<br>Democratise multilateral decision-making for FTA governance<br>Strengthen equity of FTA negotiations<br>Ensure equitable capacity for policy analysis | **Strategic plan**<br>○ Redefine trade as a tool for addressing planetary health and social determinants of health<br>○ Address the need for repeated use of short-term technical and complex to implement fixes for systemic problems<br>○ Develop enabling mechanisms to ensure trade strategies can be a tool to achieve SDG 3<br>○ Consider wider application of lessons from analysis of trade related barriers to vaccine equity<br>**Multilateral decision-making**<br>○ Involve all UN recognised states in global trade governance mechanisms with clear roles and responsibilities<br>○ Create a system of general agreement and majority voting rather than requirement for unanimous support before consensus declared<br>○ Enable nations to act without fear of sanctions that limit policy space for health<br>○ Centre human rights-based approaches and discriminated voices in designing more equitable policy and decision-making processes<br>○ Develop legal requirement to fulfil extra-territorial responsibilities in the present, recognising debt justice and the need to incorporate historical reparations for colonial activity, and subsequent inequitable and welfare-punitive material and immaterial flows of goods and services<br>**FTA negotiations**<br>○ Enable prioritisation of planetary health equity<br>○ Evidence sub-national community representation in FTA negotiations<br>○ Optimise benefits and mitigate adverse impact of FTAs on LMICs essential infrastructure and resources avoiding financial cliff-edges<br>○ Create fully supported transparent and globally equitable trade negotiation and mediation systems with LMIC leadership<br>**Policy analysis**<br>○ Reduce the resource intensive nature of policy review and analysis, making it possible for LMIC countries and institutions to undertake independently without relying on discretionary access to Global North funding | **Strategic plan**<br>○ Strengthen WHO capacities to engage with and provide technical assistance on trade- and health equity -related questions<br>○ Convene joint working programme led by WHO, bringing World Health Assembly participants and observers into conversation with WTO and WIPO to measure policy gaps against priorities for vaccine equity held by all nations<br>○ Require joint working for next round of pandemic treaty negotiations<br>○ Bring technical mitigations against vaccine inequity and incremental technical improvements into one workstream<br>○ Map steps required to ensure trade strategies can be a tool to achieve SDG 3<br>○ Translate analytical framework for application to other public health problems<br>**Multilateral decision-making**<br>○ Formally agree upon and prioritise the Determinants of Vaccine Equity (DVE) in decision-making on international trade policy<br>○ Strengthen role of human rights in decision-making and interpretation of trade and investment agreements<br>○ Require comprehensive health impact assessment of new and revised FTAs and associated policies<br>○ Provide an independent voice to advocate for non-WTO member states and people of disputed territories<br>**FTA negotiations**<br>○ Exclude essential health services like immunisation from FTAs<br>○ Require equity impact assessment in advance of FTA development<br>○ Establish a programme of engagement and joint work with discriminated communities so that equity is designed into future negotiations and revisions<br>○ Ensure that representative public health voices are present in all FTA negotiations<br>○ Provide formal observer status for FTA negotiations by national public health bodies and civil society groups<br>○ Establish a programme to monitor and address power imbalances in FTA negotiations, defining delegate numbers, testing, and evaluating ways of working to optimise global representation<br>○ Convert best endeavour agreements in health and environmental protection clauses and side letters into enforceable legislative requirements that hold corporations to account<br>**Policy analysis**<br>○ Enable transparency and access to literature and public data globally<br>○ Include vaccine availability, access, and equity in assessment of how trade agreements relate to policy space for health policies and health systems financing<br>○ Support automation of processes of finding, identifying and prioritising literature for review to maximise the use of scarce expert resources, including through natural language processing<br>○ Require search engines and repositories approved for use in literature review in policymaking, research, and teaching to include access to published research and policy documents from the Global South, particularly in-country NGOs and civil society organisations working with stigmatised and minoritised populations |
| **Pre-commercialisation**<br>(Fig 2, 1-2)<br>Address global research inequity | ○ Definition of research goals by and with LMIC stakeholders<br>○ Routine sharing of knowledge and know-how to enable globally equitable design and scale up of vaccine programmes | ○ Require corporate bodies to fulfil public tasks as a condition of public funding of research, including funding in kind e.g. use of health facilities or human volunteers<br>○ Enhance clinical trial transparency and assessment of cost-effectiveness of novel treatments against existing medicines, including new vaccines<br>○ Reduce the scale and duration of intervention generated inequity by ensuring that novel health interventions can be implemented in LMIC populations as a priority<br>○ Ensure full sharing of knowledge and know-how regarding use of vaccine components including any technological innovations<br>○ Maximise distributive potential during health emergencies as an obligation for companies and other parties commercialising research |
| **Commercialisation**<br>(Fig 2, 3)<br>Establish mechanisms to strengthen the global IP pool | ○ Make TRIPS+ waivers easily enforceable in emergency scenarios<br>○ Strengthen global IP pool to allow essential technologies and platforms to be safely produced in and for LMICs | ○ Create working group to prioritise transition to more inclusive global IP pool, built around existing endeavours of WHO with WIPO support<br>○ Reduce and geographically limit exclusive licensing practices to a level compatible with ensuring compliance with SDG 3<br>○ Create legislation to stop anti-competitive patenting behaviours (patent thickets and evergreening are prominent examples) that prevent timely patent challenges and generic production necessary for access<br>○ Expand list of essential technologies which cannot be licensed exclusively<br>○ Enshrine legal requirement for equitable access at research translation rather than procurement stage |
| **Procurement**<br>(Fig 2, 4-7)<br>Implement more equitable models of global financing and procurement | ○ Democratise finance policymaking and debate mechanisms<br>○ Strengthened investment accountability to support sustainable health interventions based on SDGs<br>○ Co-create adaptable, inequity-focused financial support models not based on Gross National Income cut-offs | ○ Collaboration on vaccines procurement to ensure production quality and sustainability<br>○ Account for need, purchasing power parity, and affordability in financial support without imposed conditions or compound interest<br>○ Develop and monitor a programme of knowledge exchange on financing models<br>○ Require full transparency of cost of goods, medicines and technologies including purchasing power parity |

### Strengths and Limitations of This Study

We examined publicly available material that documented and analysed existing and proposed policy positions and mechanisms. We included international policy and advocacy organisations advising or negotiating trade-related agreements, or proposing solutions to address public health in FTAs. By reviewing complementary sources on a timeline designed to analyse progress towards the SDGs, particularly universal access to vaccines, we achieved saturation of key themes [85]. However, we could not identify all potential stakeholders due to gaps in discoverability, including global representation in on-line databases, language restrictions, and a Westernised lens on free trade in multilateral organisations [86, 87]. This research was designed to connect what is currently recommended and find the gaps, rather than establish new evidence of causal relationships. We recognise that, as Pandemic Agreement negotiations have developed, additional evidence is emerging. Our findings, therefore, must be considered as the minimum required for action and we are conscious that novel approaches, alternative narratives, and priorities for action from those populations most affected by the adverse impact of trade-related factors on vaccine equity may have been overlooked or misinterpreted [34].

### Towards a New Framework

We found that action to address vaccine inequity could be evaluated using the 3R framework. By taking a systems approach, the relationships between specific Technical, Collaborative, and Determinants interventions were mapped onto Meadows’ points of leverage to intervene in a system, highlighting their transformative potential [36]. Achieving vaccine equity requires action on two fronts: a strategic plan bringing together the implementation of incremental and transformational improvements and a broader framework that centres DVE.

The systems map of factors affecting vaccine equity shows the interlinked nature of the action required. Technical recommendations, for example, depend on new forms of collaboration by addressing areas where policies affecting the right to health are contested. Without shifts to the wider context in which technocratic measures evolve, access initiatives remain reactive, politically unfeasible, at risk of capture or overwhelm by corporate interests as with COVAX [88]. For example, compulsory licensing and/or waiving trade secrets (Technical) to enhance production of and access to vaccines are necessary due to a lack of equity in research and technology transfer (Determinants), as seen with SARS CoV-2 vaccines [1], but even pooling mechanisms (Collaborative) are not employed, reflecting fear of backlash, sanction or non-preference in FTAs (Table 1, b.iii.1).

While development, application and evaluation of technical fixes can mitigate harm, these measures alone will not achieve vaccine equity. For example, where the policy literature focused on tariff reductions to lubricate the production chain (Table 1, f.), FTAs could, instead, exclude essential health services such as immunisation, with vaccines as essential medicines excluded or technically exempted from the articles on procurement, investment and commercialisation of services that contribute to inequities in access [1]. A framework for addressing vaccine inequity must prioritise the Determinants of health, while developing new policy spaces by strengthening Collaborative Mechanisms to make changes stick, and then applying Technical Mechanisms to enable implementation. Pandemic Agreement negotiations could still provide the basic wiring with the Conference of Parties and supporting infrastructure as fora for such measures.

### Addressing the Determinants of Vaccine Equity (DVE)

The vaccine requirements of populations with high exposure and risk of harm during the COVID-19 pandemic could have been predicted and mitigated if the DVE had been considered, Technical and Collaborative mechanisms aligned and the amenable nature of the impact of the pandemic on planetary health considered. Instead, countries with high-risk environments and significant levels of multimorbidity, including Global South nations that hosted clinical trials, like South Africa [89], experienced avoidable harm from delayed supply and excess cost of vaccines [90]. Few recommendations supported policy action to manage countries’ evolving health needs and inequities, which will be exacerbated and magnified by planetary crises. Precipitous GDP-related removal of support when reaching externally imposed thresholds was also hardly covered. Global actors responsible for vaccine programmes must acknowledge FTA-related factors and protect against increasing health inequities, rather than presuming increasing national income as result of trade will enable universal access to healthcare. This is an example of a high leverage change that cannot be addressed as an easy institutional reform or technical package because it requires a more fundamental shift in approach [37]. While there is a requirement to enact the existing evidence on implementing change more comprehensively, there is a broader requirement to address insights from foresight and emergence studies to increase readiness to pre-empt crises [91]. This is evident from the poor progress towards most of the 169 SDG targets [25], which our analysis of policy recommendations begins to explain with reference to SDG 3. Where potentially transformational recommendations were found in the literature, they were not framed comprehensively or strategically. Transforming complex systems such as international trade requires more than discrete interventions. A systems approach is necessary, mapping out the population and planetary impacts of current and potential interventions, uncovering key relationships and emerging dynamics, exploring multiple perspectives and timeframes [92, 35, 93]. Transformation can therefore be seen as multiple interventions acting synergistically [94], identifying and assembling building blocks for change [91],Here, this would include changes in multilateral decision-making processes, opening up discussion of actions that would prioritise and protect space for health equity.

### Summary of Overarching Recommendations for Multilateral Decision-Making (Table 2)

Current WTO consensus processes allow HICs to block equitable actions. A system of general agreement and majority voting would enable global majority governments to have greater say. This must be coupled with deep reflection on coloniality within multilateral institutions, and outreach to marginalised, discriminated, and working-class populations at disproportionate risk, who can guide rights-based decision making. Legislative space must be created for countries to act without fear of sanctions that limit health-related policy space. Addressing DVE requires, at minimum, health and equity impact assessment of FTAs and related policies, coupled with a critical systems view. This also means recognising debt justice and agreeing reparations for the colonial and extractive practices associated with vaccine dependency [95]. Table 2 juxtaposes these changes to other overarching recommendations, and presents immediate steps towards their actualisation.a

### Building Blocks

The WTO and WHO now have Global South leadership and more progressive ambition than before the pandemic, which offers new hope. Global negotiations to develop a pandemic treaty endeavour to address equity, trade- and IP–related issues, but they have made limited progress and risk removing effective recommendations. WTO decision-making needs to adapt to address planetary health challenges. Progress towards longer-term constitutional change and addressing wider CDH is glacial. The roles of the WTO and multilateral organisations in FTAs have been widely criticised by LMICs, especially the difficult and inequitable dispute mechanisms [96,97]. It should be possible for Member States to support strengthening the role of WHO in relation to the wider determinants of health, including planetary health, and reposition the WTO with more effective global oversight.

Multilateral bodies should have the capacity to create the conditions for countries to pass laws that hold corporations accountable for fulfilling their public responsibilities, promoting more equitable decision-making. Collective efforts should enable countries to translate currently unenforceable best endeavours agreements regarding health and its determinants into laws to protect public health, with the precautionary principle at the heart of pandemic preparedness. As a first step, this means WTO engaging with all populations regardless of UN state classification, rather than WTO members only, with space for an independent voice to advocate for peoples of disputed territories. Recent progress on multilateral governance in relation to tax provides a model worthy of further exploration as similar agreements could set out agreed minimum standards for countries to address gaps in current laws [98,99]. Meanwhile, to increase transparency and accessibility, wider access to negotiations in WTO and at national level should be given to public health bodies and formal public health associations. This could be complemented by research and background work, including with affected populations, to examine where trade and health priorities conflict or a change is required to address health.

Our results build on prior evidence of the ineffectiveness of public health safeguards in FTAs. But these cannot be seen in isolation. The ineffectiveness has been magnified by trade-related power imbalances which in turn are creating threats of public health concern, for example the emergence of novel diseases in LMICs due to deforestation and agro-industrial expansion [17]. We have not addressed these wider threats to planetary health in this paper. However, a systems approach to multilateral governance that centres DVE is a cross-sectional starting point to develop nuanced needs-based design of trade-related policy measures and analyse levers for change that reduce the emergence and severity of novel diseases of pandemic potential.

## Conclusion

The complex web of policy decisions that constitute FTAs has shaped vaccine inequity and the course of the COVID-19 pandemic. There can be no international tolerance for this scale of inequity. Here we have illuminated trade as a CDH, a link previously difficult to track but made clear by analysing barriers to vaccine equity. We have shown why institutional change is often refractory, making visible the distortion of public benefits by corporate policy capture, and the prevention of transformation from sole focus on technical measures. Known injustices and harms have deepened as a result. Our framework is transferable to other public health problems, for example, environmental change and pandemic propensity.

A framework for the transformation of FTAs is urgent, with interventions developed, tested and their impact evaluated. To facilitate action and analysis, a new multilateralism is needed. Our review identified steps towards a new framework, but our methodology is limited by potential publication bias, the lack of Global South and independent community representation. Future work must reduce inequity in discoverability of scholarship and research with an easily accessed and updated policy bank for LMIC sources. Sustainable vaccine equity requires that we transform the relationship between trade and the determinants of health. This requires an overhaul of the processes by which policy is made and governed, changing how we move towards collective planetary outcomes.

## Data Availability

All databases produced in the present work are contained in the manuscript and supplementary file. Data synthesis beyond the manuscript is available upon reasonable request to the authors.

## Acknowledgements

We are grateful for the support of Dr Siddharth Basetti, NHS Scotland, for fruitful discussions and support with organising and checking data.

