## Supplementary Appendix for "Transforming Trade for Vaccine Equity: Policy Gaps and Barriers"

**Appendix 1:** description of organisations from which sources were drawn [see included document: Unformatted Tables VE]

| **Organisation** | **Type** | **Role** | **Documents** |
| --- | --- | --- | --- |
| UN Conference on Trade and Development (UNCTAD) | International multilateral organisation | “Supporting developing countries to access benefits of a globalised society.” | - Policy guides & tools - Market summaries - Online course handbook |
| World Trade Organisation (WTO) | International organisation run by member governments | “Operates a global system of trade rules, acts as a forum for negotiating trade agreements, settles trade disputes between its members and supports the needs of developing countries.” | - Trade reports - Trade obstruction lists - Policy guides - Policy summaries - Rules and protocols - Meeting memoranda - Books and chapters |
| OECD | International organisation | “Evidence-based international standards [solutions, and policies for] a range of social, economic and environmental challenges.” | - Policy advisories - Policy summaries - Trade reports - Conference proceedings - Economic assessments |
| European Union (EU) | Union of nations    Trade bloc | “Uphold and promote member nations values and interests,” “enhance economic cohesion and solidarity” among them “and promote fair and free trade” for “sustainable development within the wider world.” | - Trade reports - Policy guides - Laws - Partnership documents |
| World Health Organisation (WHO) | International organisation    United Nations agency | “Leads global efforts to expand universal health coverage,” directing and coordinating emergency responses, and “promoting healthier lives […] guided by science”. | - Policy guides - Roadmaps, reference guides and systematic reviews - National, regional, and global strategy documents - Regional consultations - Commission reports Director-General reports - Legal recommendations - Books and chapters |
| World Intellectual Property Organisation | Multilateral Organisation | “Global forum for intellectual property policy, services, information and cooperation.” | - FTA briefings - FTA negotiations |
| World Bank | Development Bank | “A family of five international organizations that make leveraged loans to developing countries” with the “twin goals of ending extreme poverty and building shared prosperity.” | - Review of trade agreements - Country partnership strategies - Pharmaceutical industry briefings |
| Other UN bodies – UNDP, UNESCAP, HDRO, IPCIG, UNU | International multilateral organisation | “Support countries in achieving the SDGs through integrated solutions.” | - Competition law report - Conference proceedings - Policy reports - Finance reports |
| African Union | Union of nations    Trade bloc | “Continental body consisting of the 55 member states that make up the countries of the African Continent.” | - International relations report - Negotiation proceedings |
| Oxfam International | Charity | “Confederation of independent non-governmental organizations came together in 1995 to share knowledge and resources and combine their efforts in the fight against poverty and injustice” | - Research papers - Trade reports - Critique of pharmaceutical industry |
| Asian Development Bank | Development Bank | Aims to eradicate extreme poverty by providing loans, assistance, grants, and equity investments, as well as facilitating policymaking. | - Trade facilitation report - Governance review |
| European Medicines Agency | Medicines Regulatory Authority | “Evaluation and supervision of medicines, for the benefit of public and animal health in the European Union (EU).” | - Pharmacovigilance reports |
| House of Commons Library | Government | “Provide a range of research and information services for members of the British Parliament.” | - Parliamentary research briefing |
| European Centre for International Political Economy (ECIPE) | Think Tank | “Independent and non-profit policy research think tank dedicated to trade policy and other international economic policy issues of importance to Europe” | - Book chapter |
| The Global Fund to Fight AIDS, Tuberculosis and Malaria | Non-profit organisation;  NGO | “Raise and invest US$4 billion a year to fight AIDS, tuberculosis and malaria”, “challenge injustice and strengthen health systems” | - Initiative - Analytical report |
| Independent Panel for Pandemic Preparedness and Response | Body of the WHO (from WHA 73.1) | Provide an “evidence-based path for the future, grounded in lessons of the present and the past to ensure countries and global institutions, including specifically WHO, effectively address health threats.” | - Legal recommendations - Background research papers |
| New Markets Lab | Policy Hackathon | “A law and development center focused on integrating economic and social considerations into the design and implementation of law and regulation” | - Policy report |
| International Centre for Research and Agroforestry | Think Tank | “Develop knowledge practices to ensure food security and environmental sustainability.” | - Book on Value Chain Development |
| South African Institute of International Affairs | Think Tank | “Independent public policy think tank advancing a well-governed, peaceful, economically sustainable and globally engaged Africa.” | - Research paper - Policy reports |
| Rajaratnam School of International Studies | Think Tank | “Strategic Studies, International Relations, International Political Economy, and Asian Studies.” | - International relations report - Reviews |
| Montreal Economic Institute | Think Tank | “[Promoting] economic liberalism education of the general public and […] efficient public policies in Quebec and Canada.” | - Policy reports |
| International Institute for Sustainable Development | Non-Governmental Organisation | “Recommends on policies regarding international trade and investment, economic policy, management of natural and social capital, and information technologies” | - Trade review of SDGs |
| ActionAid Australia | NGO | “We empower women on the frontlines of injustice to work together and transform their communities.” | - Research paper |
| Wilson Centre | Think Tank | “Non-partisan policy forum [chartered by the US congress] for tackling global issues through independent research and open dialogue to inform actionable ideas.” | - Trade outlook report |
| European Economic and Social Committee | Multilateral Organisation | “Contributes to strengthening the democratic legitimacy and effectiveness of the European Union by enabling civil society organisations from the Member States to express their views at European level” | - Report - Impact assessments |
| Asian-Pacific Economic Cooperation | Multilateral Organisation | “Inter-governmental forum for 21 member economies in the Pacific Rim that promotes free trade throughout the Asia-Pacific region.” | - Policy report |
| Institute for Democracy and Economic Affairs | Think Tank | “Research institute dedicated to promoting solutions to public policy challenges.” | - Policy report |
| Institute of Economic Affairs | Think Tank | “Improve public understanding of the fundamental institutions of a free society, with particular reference to the role of markets in resolving economic and social problems.” | - Trade briefings |
| National Bureau of Economic Research | Think Tank | “Private, nonpartisan organization that facilitates cutting-edge investigation and analysis of major economic issues.” | - Pandemic preparedness report |
| Peterson Institute for International Economics | Think Tank | “Private, nonprofit, nonpartisan research institution devoted to the study of international economic policy.” | - Trade report |
| European Council on Foreign Relations | Think Tank | “Independent research on European foreign and security policy.” | - Power Atlas |
| OFSE (Austrian Foundation for Development Research) | Think Tank | “Research centre on questions of development policy and development cooperation.” | - Summary of existing access initiatives |

**Appendix 2:** References of stakeholder documents retrieved from search carried out up to 31/03/2022 and detailed in Appendix 3.

1 United Nations Conference on Trade and Development. Trading Into Sustainable Development: Trade, Market Access, and the Sustainable Development Goals. *UNCTAD.* 2016.
2 World Health Organization. Access to Medicines and Vaccines: Implications of Intellectual Property Protection and Trade Agreements. *WHO Regional Office for South-East Asia.* 2005.
3 World Health Organization. Public health related TRIPS-plus provisions in bilateral trade agreements: a policy guide for negotiators and implementers in the WHO Eastern Mediterranean Region. *WHO Regional Office for the Eastern Mediterranean.* 2010.
4 World Health Organization. Regional strategy to improve access to medicines and vaccines in the Eastern Mediterranean, 2020–2030, including lessons from the COVID-19 pandemic. *WHO Regional Office for the Eastern Mediterranean.* 2020.
5 World Health Organization. Roadmap for access to medicines, vaccines and health product 2019-2023: comprehensive support for access to medicines, vaccines and other health products. *WHO*. 2015.
6 World Health Assembly, 72. Access to medicines and vaccines: report by the Director- General. *WHO.* 2019.
7 World Health Organization Executive Board, 142. Addressing the global shortage of, and access to, medicines and vaccines: report by the Director-General. *WHO.* 2018.
8 World Health Organization Regional Committee for Africa, 68. Roadmap for access 2019-2023: Comprehensive support for access to medicines and vaccines. *WHO Regional Office for Africa*. 2018.
9 World Health Organization. Promoting access to medical technologies and innovation, second edition: intersections between public health, intellectual property and trade: updated extract: integrated health, trade and IP approach to respond to the COVID-19 pandemic, 30 August 2021. *WHO.* 2021.
10 World Health Organization. Trade and health: building a national strategy. *WHO*. 2015.
11 World Health Organization. International trade and health: a reference guide. *WHO Regional Office for South-East Asia.* 2009.
12 World Health Organization. SEA/RC57/Inf.5 - Globalization, trade, intellectual property rights (‎IPR)‎ and health. *WHO Regional Office for South-East Asia*. ‎2004‎.
13 World Health Organization.  Promoting cooperation for regulation in trade of medical products: Report of the regional meeting, WHO-SEARO, New Delhi, India 22-24 September 2015. *WHO Regional Office for South-East Asia.* ‎2016‎.
14 World Health Organization. Address by Dr Hussein A. Gezairy Regional Director, WHO Eastern Mediterranean Region, to the regional consultation on the impact of WTO agreements: technical barriers to trade, safeguards, and anti-dumping on public health, Dubai, United Arab Emirates, 18-21 December 2004. *WHO Regional Office for the Eastern Mediterranean.* 2004.
15 World Health Organization. Strengthening legal frameworks for health in the Sustainable Development Goals. Regional Committee for the Western Pacific, 069. *WHO Regional Office for the Western Pacific. ‎*2018‎.
16 Suthar AB, Allen LG, Cifuentes S, Dye C and Nagata JM. Lessons learnt from implementation of the International Health Regulations: a systematic review. Bulletin of the World Health Organization, 96 (‎2)‎, 110 - 121E. *WHO.* 2018.
17 World Health Organization. Build back fairer: achieving health equity in the Eastern Mediterranean Region: report of the commission on social determinants of health in the Eastern Mediterranean Region. *World Health Organization Regional Office for the Eastern Mediterranean.* 2021.
18 The Organisation for Economic Co-operation and Development, and European Commission, Joint Research Centre. Understanding the Spillovers and Transboundary Impacts of Public Policies: Implementing the 2030 Agenda for More Resilient Societies. *OECD Publishing*. 2021.
19 The Organisation for Economic Co-operation and Development, and World Trade Organization. Aid for Trade at a Glance 2015: Reducing Trade Costs for Inclusive, Sustainable Growth. *OECD Publishing*. 2015.
20 The Organisation for Economic Co-operation and Development. Structural policies to deliver a stronger, more resilient, equitable and sustainable COVID-19 recovery, in Economic Policy Reforms 2021: Going for Growth: Shaping a Vibrant Recovery. *OECD Publishing*. 2021.
21 The Organisation for Economic Co-operation and Development. OECD Science, Technology and Innovation Outlook 2021: Times of Crisis and Opportunity. *OECD Publishing*. 2021.
22 The Organisation for Economic Co-operation and Development. General Assessment of the Macroeconomic situation. Chapter in: OECD Economic Outlook, Volume 2021 Issue 1. *OECD Publishing.* 2021: 11-54.
23 The Organisation for Economic Co-operation and Development. Key Policy Insights. Chapter in: OECD Economic Surveys: Denmark 2021. *OECD Publishing*. 2021: 13-49.
24 World Trade Organization. Covid-19 vaccine production and tariffs on vaccine inputs. COVID-19 Reports. *WTO*. 2021.
25 World Trade Organization. The global race to vaccinate. Economic research and trade policy analysis. *WTO*. 2021.
26 World Trade Organization. Developing & Delivering COVID-19 Vaccines Around the World: a checklist of Issues with Trade Impact. COVID-19 Reports. *WTO*. 2020.
27 World Trade Organization. Trade and Trade-Related Policy Developments. Report on G20 Trade Measures. *WTO*. 2020: 24-89.
28 World Trade Organization. World Trade Report 2021: Economic Resilience and Trade. WTO. 2021.
29 World Trade Organization. Shifting Patterns in Trade. World Trade Statistical Review 2020. *WTO*. 2020: 32-59.
30 World Trade Organization. Trade Issues Affecting Disaster Response. WTO Working Papers. *WTO*. 2017.
31 World Trade Organization. Report on G20 Trade Measures: Mid-October 2018 to mid-May 2019
32 Oshikawa M, Anaedu U and Chemutai V. Trade Policy Trends in Africa: Empirical Evidence from Twenty Years of WTO Trade Policy Reviews. African Perspectives on Trade and the WTO. *WTO*. 2016: 13.
33 World Trade Organization. Boosting trade opportunities for least-developed countries. *WTO*. 2022.
34 World Trade Organization. When Bad Trade Policy Costs Human Lives: Tariffs on Mosquito Nets. WTO Working Papers. *WTO*. 2017.
35 World Trade Organization. Indicative list of trade-related bottlenecks and trade-facilitating measures on critical products to combat COVID-19. COVID-19 Reports. *WTO*. 2021.
36 World Trade Organization. Trade in Medical Goods in the Context of Tackling COVID-19. COVID-19 Reports. *WTO*. 2020.
37 World Trade Organization. An integrated health, trade and IP approach to respond to the COVID-19 pandemic. Promoting Access to Medical Technologies and Innovation, 2nd Edition. *WTO*. 2020: 7-15.
38 World Trade Organization. Promoting Access to Medical Technologies and Innovation, 2^nd^ Edition. *WTO*. 2020.
39 Lekic M and Osakwe C. WTO Rules, Accession Protocols and Mega-Regionals: Complementarity and Governance in the Rules-Based Global Economy. Trade Multilateralism in the 21^st^ Century. *WTO*. 2017: 8
40 Davies R, Kozul-Wright R, Banga R, Capaldo J, and Gallogly-Swan K. Reforming the International Trading System for Recovery, Resilience and Inclusive Development. Division on Globalization and Development Strategies. *UNCTAD*. 2021.
41 Peters R and Prabhakar D. Export restrictions do not help fight COVID-19. News. *UNCTAD*. 2021. Available from: <https://unctad.org/news/export-restrictions-do-not-help-fight-covid-19>
42 Kuhlmann K. Handbook on Provisions and Options for Trade in Times of Crisis and Pandemic. Online: *United Nations Economic and Social Commission for Asia and the Pacific*. *UNCTAD*. 2021.
43 United Nations Conference on Trade and Development. Toolbox for Policy Coherence in Access to Medicines and Local Pharmaceutical Production. *United Nations*. 2017.
44 Lee S, Prabhakar D. COVID-19 Non-Tariff Measures: The Good and the Bad, through a Sustainable Development Lens. *UNCTAD*. 2021.
45 Erixon F, Guinea O, van der Marel E, Philipp L. The Benefits of Intellectual Property Rights in EU Free Trade Agreements. Trade and IP. *European Centre for International Political Economy*. 2022.
46 Kaberuka, PLD. The Equitable Access Initiative Report. Gavi, the Vaccine Alliance, The Global Fund, UNAIDS, UNICEF, UNDP, UNITAID, UNFPA, WHO, and the World Bank. *The* *World Bank Group.* 2016.
47 t’Hoen E, Garrison C, Boulet P, Mara K, Perehudoff K. Scaling-up Vaccine Production Capacity: Legal Challenges and Recommendations. Background Paper 6. *The Independent Panel for Pandemic Preparedness and Response*. 2021.
48 Kuhlann K, Francis T, Thomas I, Le Graet M, Rahman M, et al. Re-conceptualizing Free Trade Agreements Through a Sustainable Development Lens. A Contribution to the Policy Hackathon on Model Provisions for Trade in Times of Crisis and Pandemic in Regional and other Trade Agreements. *New Markets Lab*. 2020.
49 United Nations Development Programme. Using competition law to promote access to medicines and related health technologies in low- and middle-income countries. *UNDP*. 2017.
50 Correa C, Velásquez G. Recent international initiatives on facilitating access to vaccines against COVID-19. COVID-19 and the Global South – Perspectives and challenges. Österreichische Entwicklungspolitik. 2021.
51 Molinuevo M, Pfister AK. Look Back to See What’s Ahead: A Review of Mega-PTAs on Services and Investment that will Shape Future Trade Agreements. Report 147060. *The World Bank Group*. 2020.
52 Hoekman B, Braga C; Robert Schuman Centre for Advanced Studies. Future of the Global Trade Order. *Publications Office of the* *European Union*. 2016.
53 Oxfam. Trading Away Access to Medicines: How the European Union's trade agenda has taken a wrong turn. Policy papers and campaign reports. *Oxfam*. 2010.
54 Mayne R. Regionalism, Bilateralism, and “TRIP Plus” Agreements: The Threat to Developing Countries. Human Development Report 2005. *Human Development Report Office*. 2005.
55 Kingah S, Ooms G. Global Constitutionalism in Global Health Governance and Regional Responses: Exploring African and Latin American Compulsory License Regimes. PRARI Working Paper 15-3. *Poverty Reduction and Regional Integration*. 2015.
56 United Nations Development Programme. Making Globalization Work for the Least Developed Countries. *UNDP*. 2015.
57 Donovan J, Stoian D, Hellin J. Value Chain Development and the Poor: Promise, delivery, and opportunities for impact at scale. International Centre for Research and Agroforestry. *Practical Action Publishing*. 2020.
58 Tigere F. The WTO and Africa: The State of Play and Key Priorities Going Forward. Policy Briefing 243. *South African Institute of International Affairs*. 2021.
59 Harris M, Manning P. International leadership by a Canada strong and free. *Montreal Economic Institute*. 2007.
60 World Bank Group. Dominican Republic - Performance and learning review of the country partnership strategy for the period FY15-FY18. Country Assistance Strategy Document. *The World Bank*. 2017.
61 International Policy Centre for Inclusive Growth. Health policy in emerging economies: innovations and challenges. *United Nations Development Programme*. 2016.
62 Tipping A, Wolfe R. Trade and Sustainable Development: Options for follow-up and review of the trade-related elements of the Post-2015 Agenda and Financing for Development. *International Institute for Sustainable Development*. 2015.
63 Anuar A, Hussain N. The Resilience of Multilateralism: From Moribund to Renewal? Multilateral Matters. S. Rajaratnam School of International Studies. *Nanyang Technological University.* 2021. Available from: <https://www.rsis.edu.sg/wp-content/uploads/2021/03/Special-Issue-MM-compressed.pdf>
64 Wilson Centre. Re-Building a Complex Partnership: The Outlook for U.S.-Mexico Relations under the Biden. *Wilson Centre*. 2021.
65 Anand PB. Discussion Paper No. 2002/110 Financing the Provision of Global Public Goods. United Nations University. *World Institute for Development Economics Research. 2002*.
66 Higelin M. Inquiry into the implications of the COVID-19 pandemic for Australia’s foreign affairs, defence and trade final. ActionAid Australia submission to the Joint Standing Committee on Foreign Affairs, Defence and Trade. *ActionAid*. 2020.
67 Faria, PE. Improving EU circular economy initiatives with stronger stakeholder input. *European Economic and Social Committee*. 2021.
68 Directorate-General for Research and Innovation; European Commission. The changing face of EU-African cooperation in science and technology: past achievements and looking ahead to the future. *Publications Office of the European Union.* 2011.
69 African Union. 6^th^ European Union - African Union summit – A joint vision for 2030. *AU Information and Communication Directorate*. 2022.
70 Asian Development Bank. Asia-Pacific Trade Facilitation Report 2021: Supply Chains of Critical Goods Amid the COVID-19 Pandemic—Disruptions, Recovery, and Resilience. Asia-Pacific Trade Facilitation Report 2021. *Asian Development Bank*. 2021.
71 Helbe M, Ali Z, Lego J. A comparison of global governance across sectors: global health, trade, and multilateral development finance. ADBI Working Paper Series No 806. *Asian Development Bank.* 2018.
72 Lee YF, Anukoonwattaka W, Taylor-Strauss H, Duva Y. Regional cooperation and integration in support of a sustainable development oriented multilateral trading system. Trade, Investment and Innovation Working Paper Series. *United Nations Economic and Social Commission for Asia and the Pacific*. 2022.
73 Braga CA, Hoekman B; Robert Schuman Centre for Advanced Studies. Future of the Global Trade Order (2^nd^ Edition). *Publications Office of the* *European Union*. 2017.
74 Stucke A, Humphreys D, Cenric DS. Improving Access and Affordability for Biopharmaceuticals in Emerging Markets: A briefing Paper from the Biopharmaceutical Think Tank Discussion at the 2019 IFC Global Private Health Care Conference. *The* *World Bank*. 2019.
75 Leonard M. The Power Atlas: Seven battlegrounds of a networked world. *European Council on Foreign Relations.* 2021.
76 General Secretariat of the Council of the European Union. The Africa-European Union strategic partnership: meeting current and future challenges together. *Publications Office of the European Union.* 2011.
77 Fowler P. Harnessing Trade for Development. Policy papers and campaign reports. Oxfam International. 2000.
78 Bloemen S, Mellema T, Bodeux L. Trading Away Access to Medicines - Revisited: How the European trade agenda continues to undermine access to medicines. Policy papers and campaign reports. Oxfam International. 2014.
79 World Health Organization. Regional Health Forum, Vol 15 No 1. *WHO Regional Office for South-East Asia.* 2011.
80 Chowdury A. Structural Transformation, LDC Graduation and the Coronavirus Disease 2019 Pandemic: Policy Options for Cambodia, Lao People’s Democratic Republic and Myanmar. Macroeconomic policy and financing for development. ESCAP Working Paper Series. *United Nations Economic and Social Commission for Asia and the Pacific.* 2021.
81 Yuhua BZ. APEC’s Bogor Goals Progress Report. APEC Policy Support Unit. *Asia-Pacific Economic Cooperation.* 2014.
82 Asian Development Bank. Managing Regional Public Goods: Cross-border Trade and Investment, Labor Migration, and Public Health: Workshop Proceedings - Part 2. Asian Development Bank. 2006.
83 ASEAN Prosperity Initiative. ASEAN Integration report 2021. Report No 6. *Institute for Democracy and Economic Affairs.* 2021.
84 Singham S. Eastern Promise: assessing the future of UK-India trade. IEA Briefing. *Institute of Economic Affairs.* 2021.
85 World Trade Organization. Public health and medical technologies: The imperative for international cooperation. Promoting Access to Medical Technologies and Innovation. *WTO*. 2013: 18-20.
86 World Trade Organization. Thoughts on How Trade, and WTO Rules, Can Contribute to the Post-2015 Development Agenda. WTO Working Papers. *WTO*. 2014.
87 World Trade Organization. TRIPS and public health: a handbook on the WTO TRIPS agreement. *WTO*. 2020: 13.
88 World Trade Organization. Other trade-related determinants for improving access, in Promoting Access to Medical Technologies and Innovation. *WTO*. 2013: 191-205.
89 World Trade Organization. The role of trade in economic resilience, in World Trade Report 2021. *WTO*. 2021: 64-121.
90 World Trade Organization. Why services trade matters. World Trade Report 2019. *WTO*. 2019: 50-81.
91 World Trade Organization. Strengthening Africa’s Capacity to Trade. *WTO*. 2021.
92 United Nations Conference on Trade and Development. IV: A more resilient multilateralism for trade and development beyond 2030. In: Transforming Trade and Development in a Fractured, Post-Pandemic World. *UNCTAD*. 2020.
93 Secretary General of the United Nations Conference on Trade and Development. Transforming trade and development in a fractured, post-pandemic world. Report of the Secretary-General of UNCTAD to the fifteenth session of the Conference. *UNCTAD*. 2020.
94 Ayoub IT. WIPO National Training Workshop on Intellectual Property for Diplomats. Negotiations of Free Trade Agreements. Republic of the Sudan. WIPO. 2007.
95 World Intellectual Property Organization. Regional and bilateral preferential trade agreements. WIPO Sub-Regional Workshop on Patent Policy and its Legislative Implementation. *WIPO*. 2013.
96 Ahuja A, Athey S, Baker A, Budish E, Castillo JC, et al. Preparing for a Pandemic: Accelerating Vaccine Availability. NBER Working Paper Series. *National Bureau of Economic Research.* 2021.
97 The Organisation for Economic Co-operation and Development. Using trade to fight COVID-19: Manufacturing and distributing vaccines. *OECD Publishing*. 2021.
98 Schott JJ, Goodman MP. Bringing supply chains back to Mexico: opportunities and obstacles. PIIE Briefing 21-4. *Peterson Institute for International Economics and/or its Licensors*. 2021.
99 United Nations Conference on Trade and Development. The many faces of inequality. UNCTAD SDG Pulse. *UNCTAD*. 2021.
100 The Organisation for Economic Co-operation and Development. Measuring transboundary impacts in the 2030 Agenda: conceptual approach and organisation. OECD Papers on Wellbeing and Inequalities. *OECD Publishing.* 2021.
101 The Organisation for Economic Co-operation and Development. Trade, global value chains and wage-income inequality. OECD Trade Policy Papers. *OECD Publishing*. 2015.
102 Sachs J, Lafortune G, Kroll C, Fuller G, Woelm F. Sustainable Development Report 2022. From Crisis to Sustainable Development: the SDGs as Roadmap to 2030 and Beyond. Sustainable Development Solutions Network. Cambridge University Press. 2022.
103 Chanda R. Trade in Health Services and Sustainable Development. ADBI Working Paper Series. *Asian Development Bank*. 2017.
104 Love J. Remuneration guidelines for non-voluntary use of a patent on medical technologies. *WHO.* 2005
105 World Trade Organization. Heads of WTO, WHO cite importance of open trade in ensuring flow of vital medical supplies. 2020 News. *WTO*. 2020.
106 World Health Organization. WHO SAGE Roadmap for prioritizing uses of COVID-19 vaccines: An approach to optimize the global impact of COVID-19 vaccines, based on public health goals, global and national equity, and vaccine access and coverage scenarios. COVID-19: Vaccines. *WHO*. 2023.
107 World Health Organization. Strategic preparedness, readiness and response plan to end the global COVID-19 emergency in 2022. COVID-19: Critical preparedness, readiness and response. *WHO*. 2022.
108 World Health Organization. Global strategy and plan of action on public health, innovation and intellectual property: Report by the Director-General. *WHO*. 2021.
109 United Nations Department of Economic and Social Affairs. UN Technology Facilitation Mechanism (TFM): Harnessing Science, Technology and Innovation to achieve the Sustainable Development Goals. United Nations. 2020.
110 Karingi SN, Pesce O, Mevel S. Preferential Trade Agreements in Africa: Lessons from the Tripartite Free Trade Agreements and an African Continent-Wide FTA. *WTO*. 2016: 14.
111 The Organisation for Economic Co-operation and Development. Deep Provisions in Regional Trade Agreements: How Multilateral-friendly? An Overview of OECD Findings. OECD Trade Policy Papers. *OECD Publishing*. 2014.
112 Bown CP. Mega-Regional Trade Agreements and the Future of the WTO. *Global Policy* 2017: **8**, 1: 107-112.
113 Bown CP. Why the World Trade Organization is Critical for Vaccine Supply Chain Resilience During a Pandemic. *World Bank Group*. 2022.
114 Webb D. Progress on UK free trade agreement negotiations. Research Briefing No 9314. House of Commons Library. *UK Parliament*. 2022.
115 World Health Organization. Middle Income Countries. Immunization Agenda 2030. *WHO*. 2020.
116 Narayanasamy S, Curtis LH, Hernandez AF, Woods CW, Adam SJ et al. Lessons from COVID-19 for pandemic preparedness: proceedings from a multistakeholder think tank. Clinical Infectious Diseases. 2023 Dec 15;77(12):1635-43.

**Appendix 3:** PRISMA diagram showing search terms, titles screened and included, and reasons for exclusion.

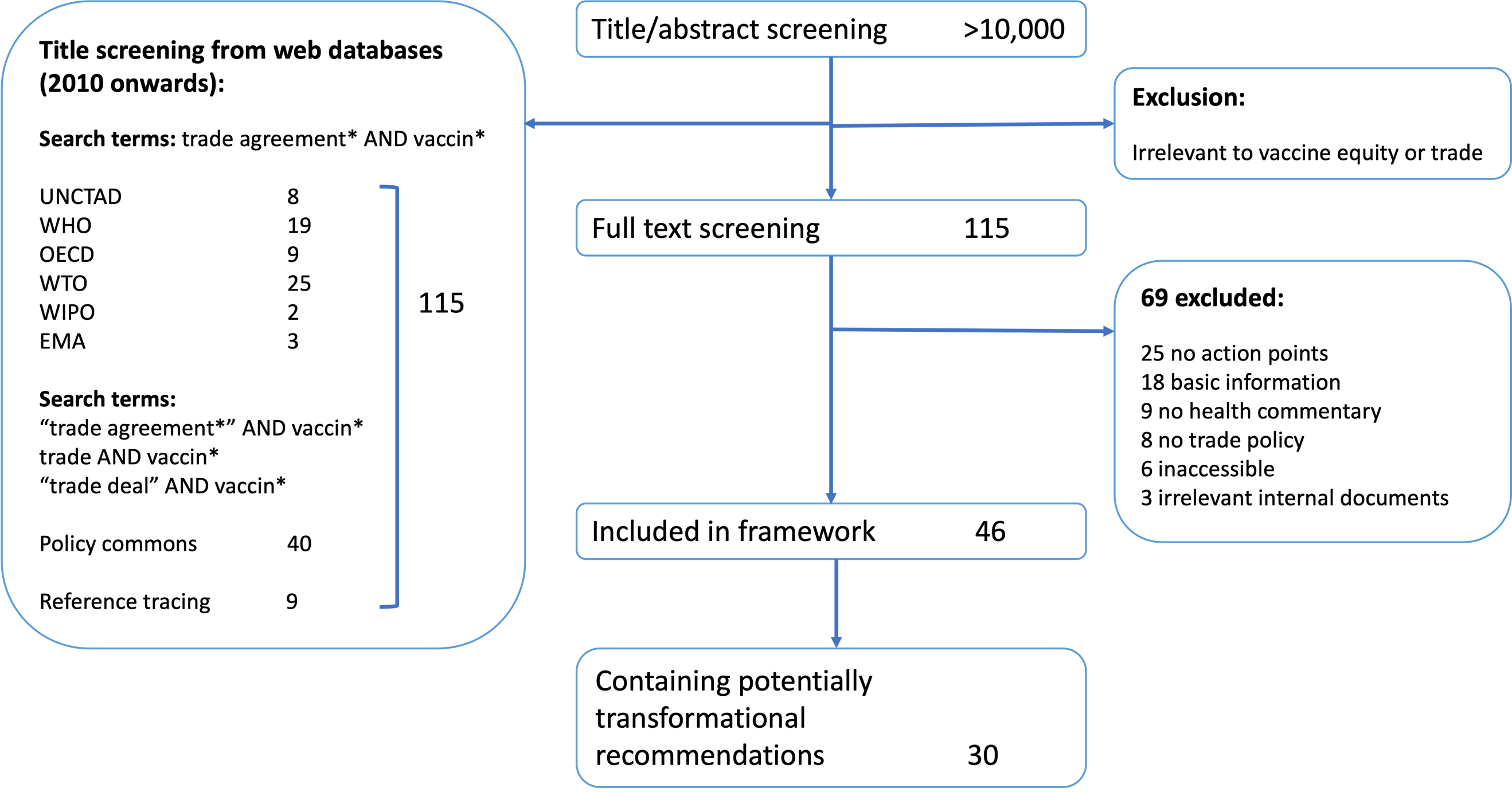
